# Multi-View Echocardiographic Embedding for Accessible AI Development

**DOI:** 10.1101/2025.08.15.25333725

**Authors:** Takeshi Tohyama, Ahram Han, Dukyong Yoon, Kenneth Paik, Brian Gow, Nura Izath, Jacques Kpodonu, Leo Anthony Celi

## Abstract

**Background and Aims:** Echocardiography serves as a cornerstone of cardiovascular diagnostics through multiple standardized imaging views. While recent AI foundation models demonstrate superior capabilities across cardiac imaging tasks, their massive computational requirements and reliance on large-scale datasets create accessibility barriers, limiting AI development to well-resourced institutions. Vector embedding approaches offer promising solutions by leveraging compact representations from original medical images for downstream applications. Furthermore, demographic fairness remains critical, as AI models may incorporate biases that confound clinically relevant features. We developed a multi-view encoder framework to address computational accessibility while investigating demographic fairness challenges.

**Methods:** We utilized the MIMIC-IV-ECHO dataset (7,169 echocardiographic studies) to develop a transformer-based multi-view encoder that aggregates view-level representations into study-level embeddings. The framework incorporated adversarial learning to suppress demographic information while maintaining clinical performance. We evaluated performance across 21 binary classification tasks encompassing echocardiographic measurements and clinical diagnoses, comparing against foundation model baselines with varying adversarial weights.

**Results:** The multi-view encoder achieved a mean improvement of 9.0 AUC points (12.0% relative improvement) across clinical tasks compared to foundation model embeddings. Performance remained robust with limited echocardiographic views compared to the conventional approach. However, adversarial learning showed limited effectiveness in reducing demographic shortcuts, with stronger weighting substantially compromising diagnostic performance.

**Conclusions:** Our framework democratizes advanced cardiac AI capabilities, enabling substantial diagnostic improvements without massive computational infrastructure. While algorithmic approaches to demographic fairness showed limitations, the multi-view encoder provides a practical pathway for broader AI adoption in cardiovascular medicine with enhanced efficiency in real-world clinical settings.

**Structured graphical abstract or graphical abstract:** *Key Question:* Can multi-view encoder frameworks achieve superior diagnostic performance compared to foundation model embeddings while reducing computational requirements and maintaining robust performance with fewer echocardiographic views for cardiac AI applications?

*Key Finding:* Multi-view encoder achieved 12.0% relative improvement (9.0 AUC points) across 21 cardiac tasks compared to foundation model baselines, with efficient 512-dimensional vector embeddings and robust performance using fewer echocardiographic views.

*Take-home Message:* Vector embedding approaches with attention-based multi-view integration significantly improve cardiac diagnostic performance while reducing computational requirements, offering a pathway toward more efficient AI implementation in clinical settings. 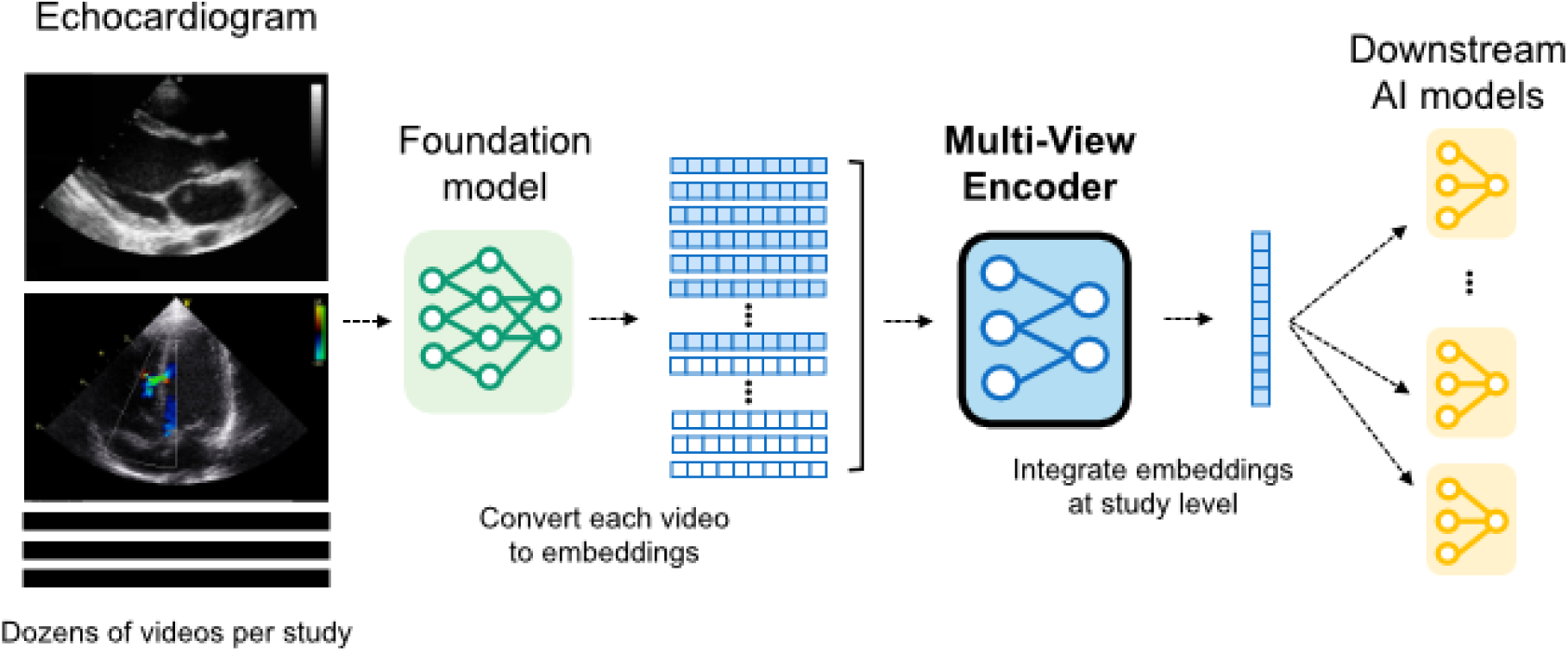

*Translational Perspective:* Our proposed multi-view encoder framework overcomes critical barriers to the widespread adoption of artificial intelligence in echocardiography. By dramatically reducing computational requirements, the multi-view encoder approach allows smaller healthcare institutions to develop sophisticated AI models locally. The framework maintains robust performance with fewer echocardiographic examinations, which addresses real-world clinical constraints where comprehensive imaging is not feasible due to patient factors or time limitations. This technology provides a practical way to democratize advanced cardiac AI capabilities, which could improve access to cardiovascular care across diverse healthcare settings while reducing dependence on proprietary datasets and massive computational resources.

## Introduction

Echocardiography serves as the cornerstone of cardiovascular diagnostics, providing real-time visualization of cardiac structure and function through multiple standardized imaging views^1^. While recent foundation models such as EchoPrime and PanEcho have demonstrated superior capabilities across diverse cardiac imaging tasks^2–5^, their massive computational requirements and reliance on large-scale proprietary datasets create substantial barriers, limiting AI development to institutions with extensive resources^6,7^.

The inherent complexity of echocardiographic assessment—requiring comprehensive multi-view evaluation and integration of diverse imaging modalities including color Doppler— further complicates these implementation challenges^8,9^. Vector embedding (VE) methods offer a promising alternative approach that can substantially reduce these barriers by leveraging representations extracted from pre-trained models for diverse downstream applications ^10,11^.

However, beyond accessibility considerations, demographic fairness remains a critical challenge in medical AI. Models may unintentionally learn spurious demographic shortcuts rather than clinically relevant pathophysiological features, thereby creating potential biases that can lead to disparate performance across demographic groups. ^12^ This concern persists when leveraging foundation model representations. While adversarial learning approaches have been proposed to mitigate such biases, their effectiveness in complex medical imaging tasks remains unclear, highlighting the intricate nature of fairness challenges in healthcare AI^13,14^.

Here, we introduce a multi-view encoder (MVE), a novel VE framework that achieves two major advances in echocardiographic AI. First, comprehensive multi-view integration where our approach leverages a masked transformer encoder to aggregate view-level representations into unified study-level embeddings, mimicking clinical practice where information is synthesized across multiple imaging planes for comprehensive cardiac assessment. Second, computational accessibility, where our VE approach extracts compact, clinically-relevant representations, dramatically reducing computational requirements while maintaining diagnostic performance and enabling broader AI adoption in resource-limited settings. Additionally, we systematically investigate the complexities of demographic fairness in medical AI through adversarial learning approaches, revealing fundamental challenges in achieving algorithmic fairness in echocardiographic analysis. This integrated framework enables healthcare institutions with limited AI resources to develop sophisticated cardiac diagnostic models locally without requiring massive computational infrastructure.

## Methods

### Study Design and Data Source

This retrospective study utilized the MIMIC-IV and MIMIC-IV-ECHO datasets^15–17^, publicly available anonymized databases of echocardiographic examinations collected at Beth Israel Deaconess Medical Center. We included all patients between 2017 and 2019 in the MIMIC-IV-ECHO dataset with clinical diagnostic information. Only echocardiographic examinations that contained both DICOM video files and corresponding measurement records were included. The final dataset included 7,169 echocardiographic studies with corresponding echo measurements and clinical diagnoses. Use of the MIMIC-IV database did not require additional institutional review board approval as the data were previously de-identified in accordance with HIPAA Safe Harbor provisions. All analyses were conducted in accordance with the Declaration of Helsinki, and this study is reported in accordance with TRIPOD+AI statement^18^. No prospective protocol registration was conducted as this retrospective analysis utilized existing publicly available datasets. No patient and public involvement was undertaken in this retrospective study.

### Outcome Definition for Clinical Tasks

We defined 21 binary classification tasks encompassing echocardiographic measurements, functional assessments, and clinical diagnoses (Table 1). Primary outcomes included echo measurements (e.g., reduced ejection fraction, right ventricular dysfunction, and valvular disease) and clinical diagnoses such as cardiomyopathy and myocardial infarction. No additional outcome assessment or re-interpretation was conducted for this study. All outcomes were dichotomized using clinically validated thresholds established in current guidelines ^19,20^. This approach enabled unified evaluation and interpretation of diagnostic performance across diverse cardiac conditions.

**Table 1:** MIMIC-IV-ECHO patient characteristics.

| Variables |  | Missing | Overall<br>7169 |
| --- | --- | --- | --- |
| Video count, median [Q1, Q3] |  | 0 | 41 [34,48] |
| Sex, n (%) | Female | 417 (5.8) | 3478 (51.5) |
|  | Male |  | 3274 (48.5) |
| Race, n (%) | White | 498 (6.9) | 4777 (71.6) |
|  | Black |  | 1232 (18.5) |
|  | Hispanic |  | 287 (4.3) |
|  | Asian |  | 179 (2.7) |
|  | Others |  | 196 (2.9) |
| Aortic Root/BSA: >2.1cm/m2, n (%) |  | 736 (10.3) | 423 (6.6) |
| LVDd > 5.8cm (male) or 5.2cm (female), n (%) |  | 519 (7.2) | 663 (10.0) |
| LAVI > 34mL/m2, n (%) |  | 2058 (28.7) | 2104 (41.2) |
| LVMI > 115g/m2 (male) or 95g/m2 (female), n (%) |  | 591 (8.2) | 3256 (49.5) |
| LVEF < 50%, n (%) |  | 1804 (25.2) | 830 (15.5) |
| Impaired RV Function, n (%) |  | 450 (6.3) | 1064 (15.8) |
| Mild AR or greater, n (%) |  | 171 (2.4) | 1491 (21.3) |
| Mild AS or greater, n (%) |  | 171 (2.4) | 726 (10.4) |
| Mild MR or greater, n (%) |  | 164 (2.3) | 3340 (47.7) |
| Mild TR or greater, n (%) |  | 168 (2.3) | 3508 (50.1) |
| Moderate AR or greater, n (%) |  | 171 (2.4) | 154 (2.2) |
| Moderate AS or greater, n (%) |  | 171 (2.4) | 413 (5.9) |
| Moderate MR or greater, n (%) |  | 164 (2.3) | 856 (12.2) |
| Moderate TR or greater, n (%) |  | 168 (2.3) | 974 (13.9) |
| TRPG > 31mmHg, n (%) |  | 1353 (18.9) | 1949 (33.5) |
| E/e' > 15, n (%) |  | 4064 (56.7) | 549 (17.7) |
| Dilated IVC, n (%) |  | 4904 (68.4) | 448 (19.8) |
| Dilated cardiomyopathy, n (%) |  | 417 (5.8) | 455 (6.7) |
| Hypertrophic cardiomyopathy, n (%) |  | 417 (5.8) | 122 (1.8) |
| Myocardial infarction, n (%) |  | 4968 (69.3) | 160 (7.3) |
| Pulmonary embolism, n (%) |  | 5189 (72.4) | 68 (3.4) |

BSA, body surface area; LVDd, left ventricular diameter (diastolic); LAVI, left atrial volume index; LVMI, left ventricular mass index; LVEF, left ventricular ejection fraction; RV, right ventricle; AR, aortic regurgitation; AS, aortic stenosis; MR, mitral regurgitation; TR, tricuspid regurgitation; TRPG, tricuspid regurgitation pressure gradient; E/e', ratio of early mitral inflow velocity to early diastolic mitral annular tissue velocity; IVC, inferior vena cava.

### VE Extraction from Foundation Model

Input predictor variables consisted of 512-dimensional VEs generated by processing echocardiographic DICOM videos through the EchoPrime foundation model, a multi-view vision-language model trained on over 12 million video-report pairs using contrastive learning^3^. The included echocardiographic studies contained a median of 41 videos per study (maximum 118) representing multiple standardized echocardiographic videos including parasternal, apical, and subcostal views. No manual feature extraction or subjective assessment of echocardiographic images was performed by the investigators.

### MVE Model Development

The model employed a transformer-based architecture with two transformer layers and eight attention heads each, optimized for computational efficiency following compact transformer approaches^21–23^. This architecture processes variable numbers of cardiac views simultaneously (Figure 1). The MVE aggregates variable numbers of cardiac view embeddings per study using attention masking mechanisms, supporting up to 128 input views. During MVE pretraining, an adversarial learning component with gradient reversal layers was added to suppress demographic predictability from the learned study-level representations.^13,24^.

**Figure 1:**
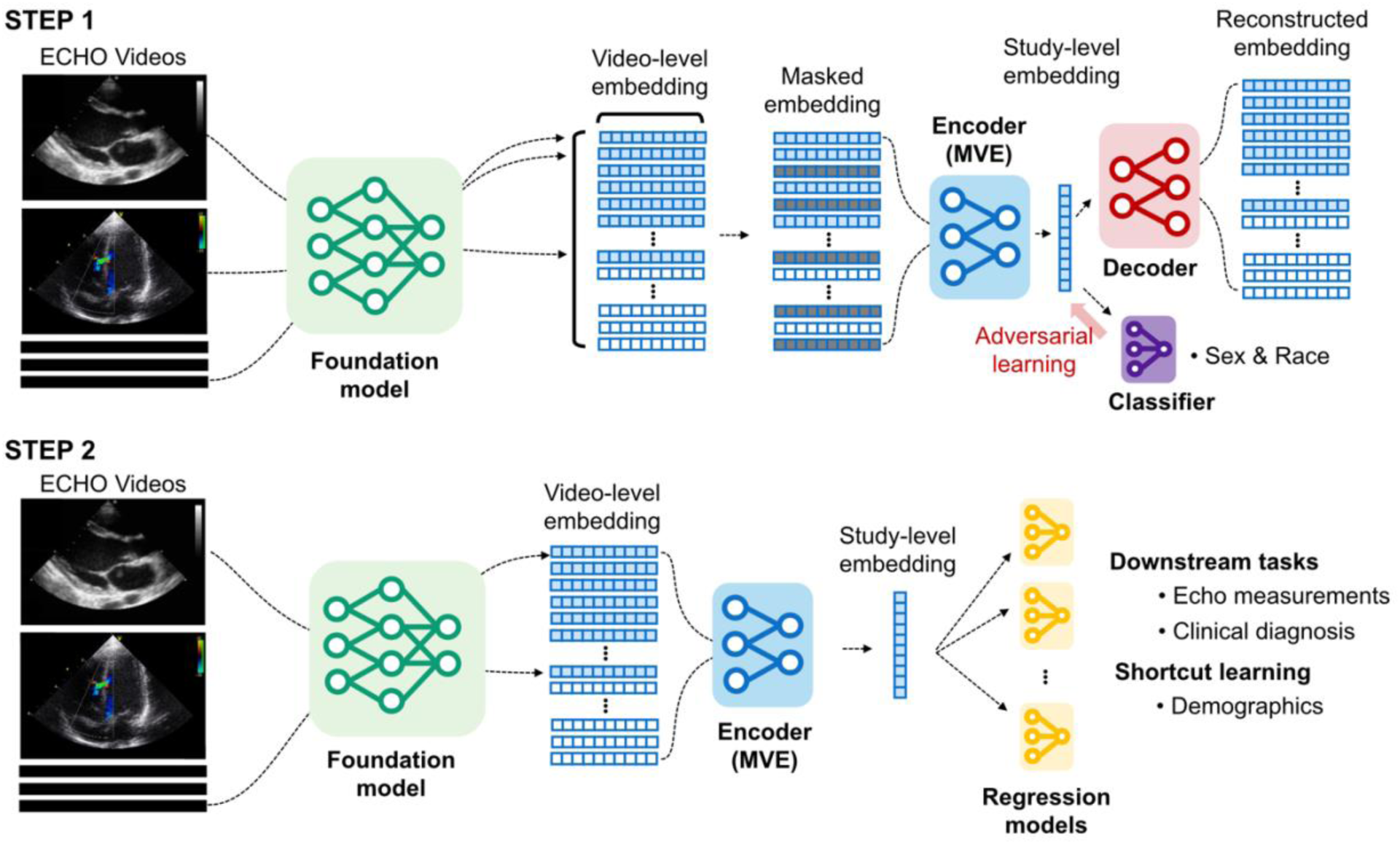
T**w**o**-step framework for multi-view echocardiographic representation learning Step 1: Pre-training for multi-view encoder (MVE).** Echocardiographic video files are processed through the foundation model to generate 512-dimensional video-level embeddings. The MVE framework uses a shared transformer with attention-based aggregation capabilities to simultaneously process multiple video-level embeddings and learn study-level embeddings through reconstruction tasks. An adversarial learning module (red) for demographic classification (sex and race) is included, penalizing the encoder for learning demographic-specific features. This fairness-aware training process aims to produce representations that maintain clinical utility while reducing the ability to predict patient demographics. **Step 2: Downstream task evaluation.** Study-level embeddings for test datasets are generated by the pre-trained MVE. These embeddings are systematically evaluated across 21 downstream clinical tasks, including echocardiographic measurements and diagnoses, and two demographic predictions to assess potential shortcut learning.

The adversarial framework included separate discriminator networks for sex prediction (3 classes: female/male/unknown) and race prediction (4 classes: white/black/other/unknown), trained simultaneously with clinical tasks. The gradient reversal layer applies negated gradients (scaled by adversarial weight ω) during backpropagation, forcing the encoder to learn representations with lower predictability of demographic attributes while maintaining performance in clinical tasks. We systematically varied the adversarial weight (ω) from 0 to 1.0 (0, 0.1, 0.2, 0.5, 1.0) to investigate trade-offs between model performance and demographic fairness. An ω of 0 indicates no adversarial learning, while ω of 1 represents strong adversarial learning.

Data were split into training (42.5%), validation (7.5%), and test (50%) sets using subject-based splitting to ensure no same patients appeared across multiple splits (Supplementary Figure 1). We split the data equally, allocating 50% for MVE training and validation and 50% for downstream task evaluation using 4-fold cross-validation. Our study consisted of two steps: (1) pre-training MVE using 50% random view masking for multi-view representation learning, and (2) evaluation with downstream tasks using VE representations extracted from MVE. The MVE was trained for 50 epochs using the AdamW optimizer with learning rate 1×10⁻⁴ and early stopping based on validation loss and architectural parameters were selected based on best practices from previous studies and preliminary validation experiments. The model architecture supports a variable number of echocardiographic views by employing attention masking mechanisms. It processes up to 128 videos and generates a 512-dimensional VE for each study.

### Downstream Task Performance Evaluation

We employed subject-based 4-fold cross-validation on the test set for downstream task evaluation (Supplementary Figure 1). This ensured no patients appeared in both training and test sets in downstream task evaluation, preventing optimistic bias from patient-specific patterns. For downstream classification tasks, we used a simple logistic regression model with L2 regularization. Primary evaluation utilized the area under the receiver operating characteristic curve (AUC). No specific class imbalance methods (e.g., resampling, cost-sensitive learning) were applied. To provide comprehensive evaluation, we supplemented AUC with accuracy and F1 scores, where the decision threshold was determined by optimizing F1 on the training data.

We compared two types of VEs for downstream task performance. Foundation VE served as the baseline, consisting of embeddings directly extracted from EchoPrime using the original data processing pipeline. This approach aggregates variable-length input sequences into fixed 15 × 512 dimensions through weighted averaging based-on view classification. MVE VEs were embeddings extracted from MVEs with varying adversarial weights, where each embedding had 512 dimensions.

### Fairness Evaluation and Impact on Clinical Task Performance

We systematically evaluated potential demographic shortcuts by training separate classifiers to predict sex and race from the Foundation VE and MVE VE representations, respectively. We employed assessment of demographic predictability as an established proxy measure for evaluating shortcut learning ^25,26^. This approach provides one of the metrics to evaluate whether learned representations have reduced reliance on demographic shortcuts.

Additionally, we assessed how adversarial learning affects actual clinical task prediction performance. To evaluate potential disparities in model performance, we compared AUC values for 21 clinical tasks across demographic subgroups (sex: female/male; race: white/black/other) and video count subgroups (<20, 20–39, 40–59, ≥60 videos per study). Furthermore, we evaluated the effects of adversarial learning weighting for four important cardiac conditions with sufficient prevalence (mild aortic stenosis, left ventricular ejection fraction <50%, impaired right ventricular function, E/e’ ratio >15) across demographic subgroups (sex: female/male, race: white/black/other).

## Statistical Analysis

The study utilized the complete available MIMIC-IV-ECHO dataset (n=7,169) for comprehensive analyses rather than conducting individual sample size calculations. This approach was justified for the following reasons: (1) the comprehensive evaluation across 21 diverse clinical tasks with varying prevalence rates (1.8% to 50.1%) would require different sample size requirements for each task, making a single calculation impractical; (2) utilizing all available data maximizes statistical power and ensures representativeness of the source population.

Performance comparison and model fairness on VEs were assessed using mixed effects models, with improvements quantified through relative improvement metrics. Detailed statistical models are provided in supplementary materials. Multiple testing correction for main performance and model fairness evaluation was performed using the Holm-Bonferroni method. All analyses were conducted using Python 3.8 with scikit-learn, PyTorch, and statsmodels.

## Results

### Dataset Characteristics

Our analysis included 7,169 echocardiographic studies with a median of 41 videos per study [interquartile range (IQR): 34-48] (Table 1). Among patients with recorded sex information (n=6,752), there were 3,478 female patients (51.5%) and 3,274 male patients (48.5%). Among patients with recorded race information (n=6,671), the racial distribution comprised white patients (71.6%, n=4,777), black patients (18.5%, n=1,232), Hispanic patients (4.3%, n=287), Asian patients (2.7%, n=179), and other racial categories (2.9%, n=196).

The echo measurements covered diverse cardiac conditions with varying prevalence, including reduced ejection fraction (LVEF < 50%: 15.5%), right ventricular dysfunction (15.8%), mild or greater mitral regurgitation (47.7%), mild or greater tricuspid regurgitation (50.1%), elevated tricuspid regurgitation pressure gradient (TRPG > 31 mmHg: 33.5%), and elevated E/e’ ratio (> 15: 17.7%). Clinical diagnoses included hypertrophic cardiomyopathy (1.8%), dilated cardiomyopathy (6.7%), myocardial infarction (7.3%), and pulmonary embolism (3.4%).

### MVE Achieved Performance Improvements

To establish a benchmark MVE performance for downstream tasks, we first evaluated MVE without adversarial learning (ω = 0) across 21 binary clinical tasks spanning valvular disease, functional assessment, hemodynamic parameters, and clinical diagnoses. The MVE framework demonstrated consistent performance, achieving a mean improvement of 8.4 AUC points (95% CI: 7.5-9.4). This represents an 11.2% relative improvement compared to the Foundation VE (Figure 2A). However, MVE VEs showed increased predictability for both sex and race, indicating that the performance gains were associated with demographic shortcuts.

**Figure 2:**
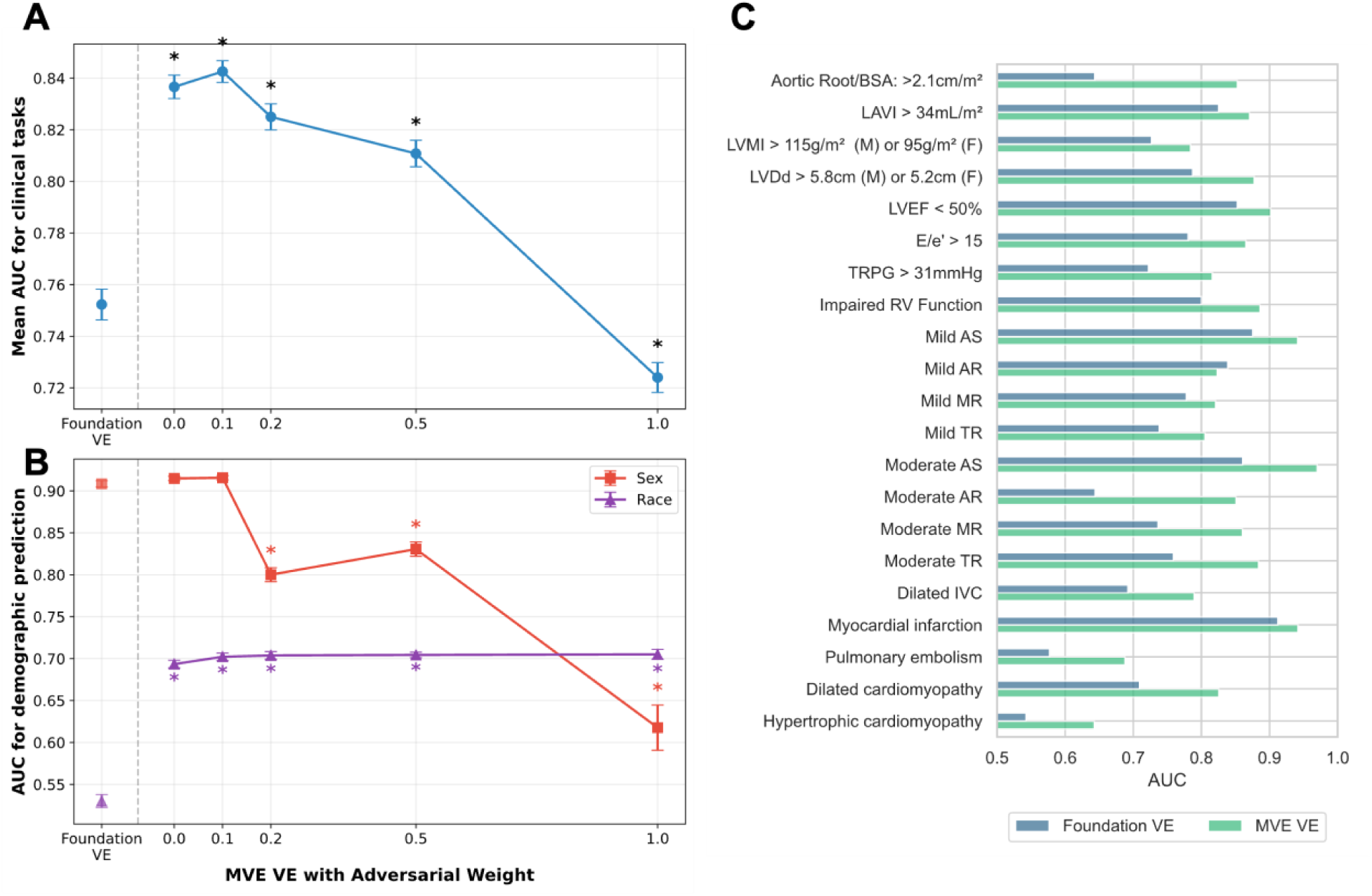
Performance and fairness trade-off analysis. **A.** Changes in area under the curve (AUC) across 21 clinical tasks for vector embeddings from multi-view encoder (MVE VE) with varying adversarial weights (ω = 0 to 1.0) compared to vector embeddings from foundation model (Foundation VE). Results represent mean ± standard error AUC across all 21 tasks from five independent experimental repeats with 4-fold cross-validation. “*” indicates statistical significance (p < 0.05) compared to Foundation VE. **B.** Relationship between demographic predictability and adversarial learning with different weights. Sex predictability (red squares) and race predictability (purple triangles) represent mean ± standard error AUC from five independent repeats with 4-fold cross-validation. “*” indicates statistical significance (p<0.05) compared to Foundation VE. **C.** Comparison of AUC performance between Foundation VE and MVE VE with adversarial training (weight = 0.1) across the 21 clinical tasks.

To mitigate the problem of demographic shortcuts, we incorporated adversarial learning and systematically varied the adversarial weight (ω) from 0.1 to 1.0 to evaluate its effect on both downstream task performance and demographic predictability (Figure 2A and 2B). The MVE with mild adversarial learning (ω = 0.1) achieved a mean improvement of 9.0 AUC points (95% CI: 8.1–10.0), representing the highest improvement (12.0% relative improvement) over the Foundation VE, with no significant change in demographic predictability compared to MVE without adversarial learning (ω = 0). Further increases in adversarial weighting reduced sex predictability to some extent; however, this reduction was accompanied by substantial performance degradation in clinical tasks that worsened as adversarial weights increased (Figure 2A). Conversely, the reduction in race predictability was not evident across the tested range of adversarial weights, suggesting limited effectiveness of the adversarial learning approach for addressing racial demographic shortcuts in echocardiographic representations.

Performance across individual clinical tasks showed AUC improvement in 20 of 21 tasks, with a median improvement of 9.0 points (IQR: 5.7–11.1) using adversarial learning with MVE (ω = 0.1) compared to Foundation VE (Figure 2C). These improvements were consistent across other metrics, with accuracy improving by a median of 2.1 points (IQR: 1.6-2.9) and F1 scores by 7.6 points (IQR: 5.0-10.4) (Supplementary Figure 2). However, F1 scores remained low for some clinical tasks, particularly those with low prevalence such as moderate or greater aortic regurgitation (prevalence 2.2%) and hypertrophic cardiomyopathy (prevalence 1.8%), despite showing F1 score improvements.

### Performance Disparities Across Demographic Groups

Performance analysis across demographic subgroups revealed that the MVE framework consistently outperformed the Foundation VE across all evaluated subgroups (Figure 3A and 3B). Subgroup analyses revealed baseline performance differences among racial groups (p=0.001), suggesting inherent racial biases. However, there was no significant model × subgroup interaction (p=0.622), indicating that MVE or adversarial learning (ω = 0.1, 0.2) did not reduce these racial disparities. Performance improvements were consistent across all demographic groups, with no differential effects of adversarial learning on subgroup biases.

**Figure 3:**
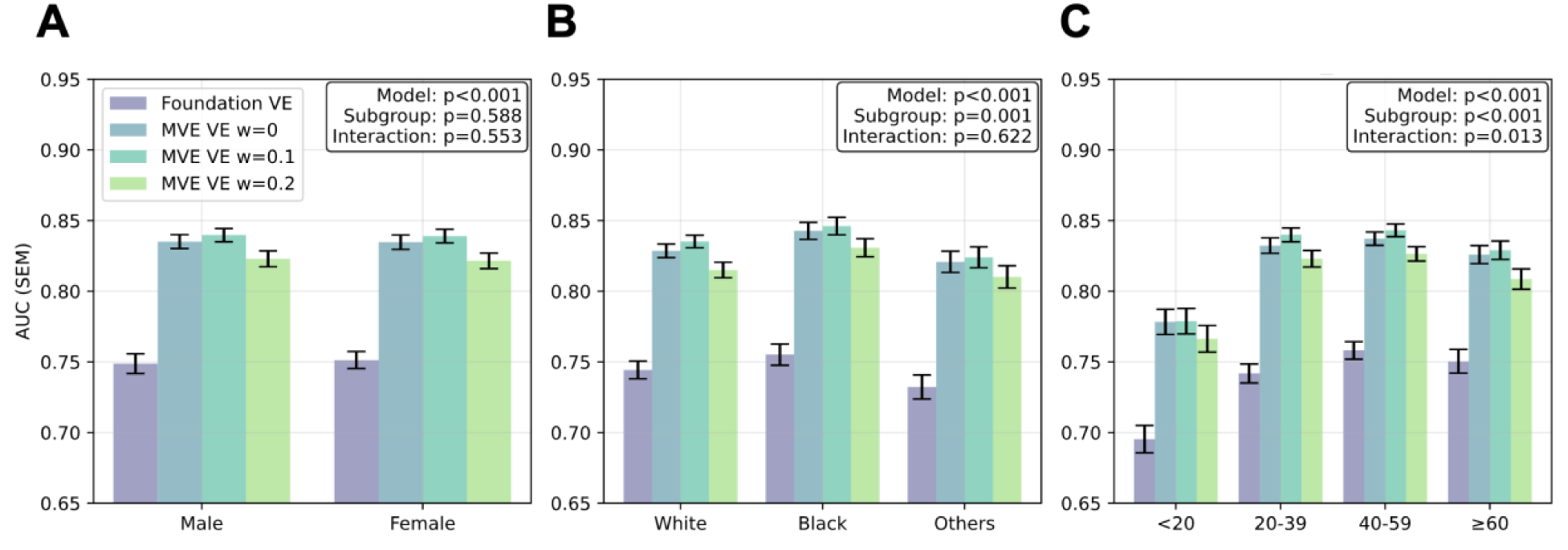
Subgroup performance analysis across demographic groups Comprehensive comparison of mean ± standard error AUC performance across 21 clinical tasks for vector embeddings from foundation model (Foundation VE) and multi-view encoder (MVE VE) stratified by demographic subgroups. Adversarial weights are shown from ω = 0 to ω = 0.2. Analysis includes stratification by sex (**A**: female vs male), race (**B**: white vs black vs other), and number of videos per study (**C**: <20, 20-39, 40-59, ≥60). P-values are provided for mixed effects models evaluating the model term, subgroup term, and model × subgroup interaction term for each comparison.

To further investigate subgroup-level performance disparities at the individual task level, we analyzed four specific cardiac outcomes (Figure 4). Among these, right ventricular (RV) dysfunction and elevated E/e′ ratio showed subtle subgroup differences. In contrast, mild aortic stenosis exhibited lower performance in the “other” race category, and reduced ejection fraction (LVEF <50%) showed higher performance in black patients. These disparities showed little improvement despite increasing adversarial weights. Indeed, strong adversarial weighting (ω ≥ 0.5) resulted in substantial performance degradation across all subgroups. Notably, clinical outcomes that exhibited baseline demographic disparities (mild AS and LVEF <50%) experienced substantial performance degradation under strong adversarial weighting (ω = 1.0). In contrast, tasks with small demographic differences at baseline (impaired RV function) were less affected by adversarial learning interventions.

**Figure 4:**
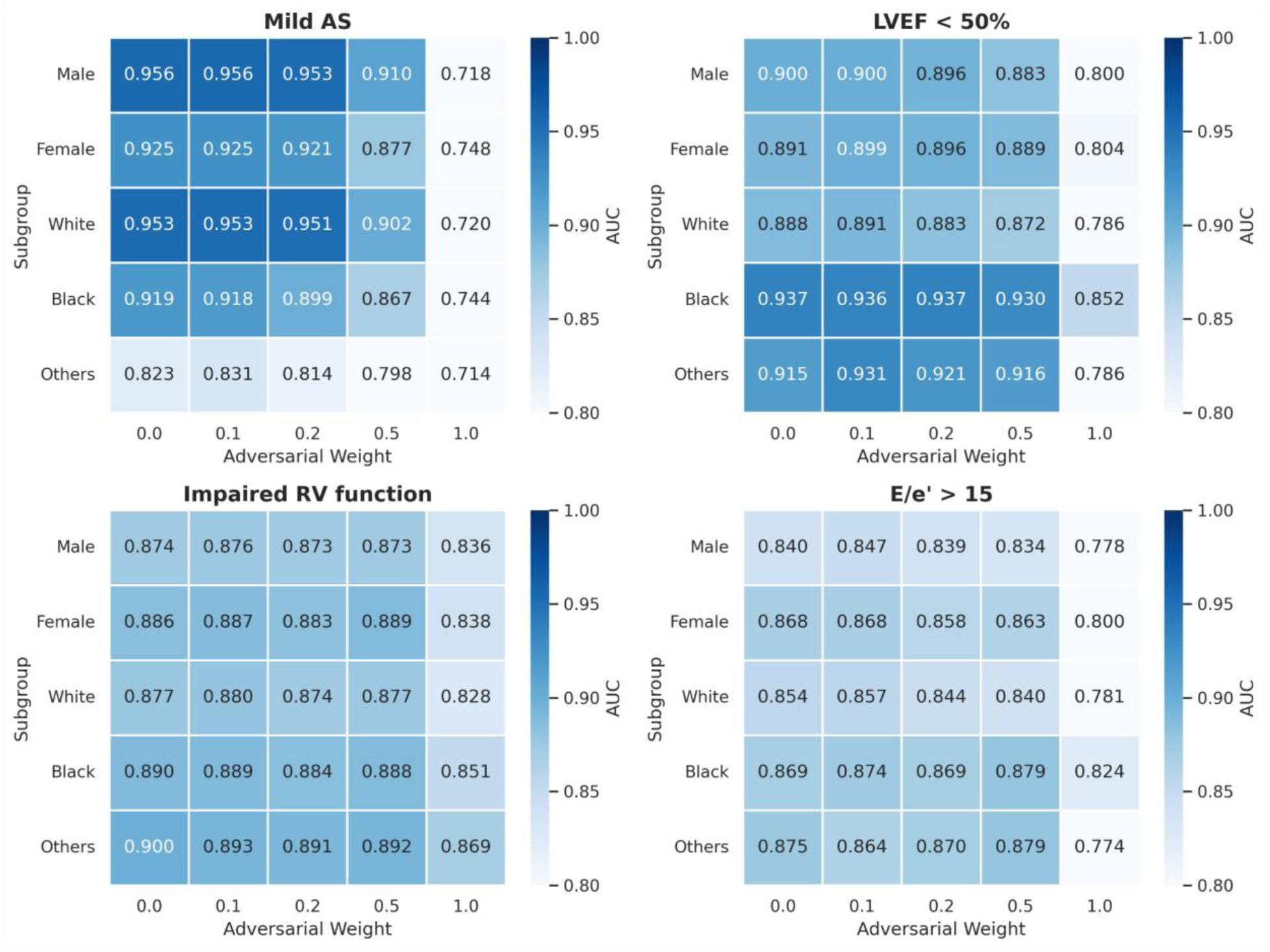
Analysis of subgroup performance differences and adversarial learning effects across demographic groups. Heatmap showing AUC performance by demographic subgroups and adversarial weighting (ω = 0-1.0) for four important clinical tasks: mild aortic stenosis (AS), reduced ejection fraction (LVEF < 50%), impaired right ventricular (RV) function, and E/e’ > 15. Color intensity represents AUC values, with darker colors indicating higher predictive performance. Each row represents different demographic subgroups (female, male, white, black, others) and each column represents increasing adversarial weights from 0 to 1.0.

### Robustness to Limited View Availability

As shown in Figure 3C, video count analysis revealed a significant subgroup difference (p<0.001), indicating that fewer echocardiographic views directly impaired downstream task performance. Additionally, a significant model × subgroup interaction (p=0.013) demonstrated that MVE and Foundation VE responded differently to varying examination completeness. Specifically, while Foundation VE performance continued to improve with increasing views up to 40-59 per study, MVE VE reached a performance plateau at 20-39 views, demonstrating efficient pathophysiological feature extraction with fewer views.

## Discussion

### Key Findings and Clinical Significance

Our study demonstrates that the multi-view encoder (MVE) with adversarial learning framework achieved substantial improvements across 21 clinical tasks, with a mean improvement of 9.0 AUC points (12.0% relative improvement) compared to foundation model baselines. These gains were particularly pronounced in tasks requiring multi-view integration and color Doppler information, such as valvular disease assessment, where comprehensive cardiac evaluation across multiple echocardiographic perspectives provides advantages over single-view analysis^5^. The consistency of improvements across diverse cardiac outcomes—from structural abnormalities to clinical diagnoses—suggests broad clinical utility for comprehensive cardiac evaluation.

A particularly significant finding was MVE VE’s enhanced robustness when fewer echocardiographic views were available. While Foundation VE showed continued dependence on increasing view numbers, MVE achieved comparable performance between studies with 20-39 views and those with over 40 views (Figure 3C). This view compensation effect has profound clinical implications, as real-world echocardiographic examinations often vary in completeness due to patient factors, time constraints, or equipment limitations. The ability to maintain performance in clinical tasks with fewer views would enable rapid screening protocols in outpatient settings and reliable performance in emergency departments where comprehensive examinations may not be feasible.

Furthermore, our VE approach addresses a fundamental challenge in medical AI: the democratization of advanced capabilities without requiring massive computational infrastructure.

MVE training completes within hours on affordable GPU hardware compared to weeks-to-months required for foundation model training on specialized infrastructure. By leveraging the MVE VE framework that aggregates echocardiographic view-level representations into study-level embeddings, smaller healthcare institutions can achieve rapid AI implementation and substantial improvements without proprietary datasets or industrial-scale resources ^10,11^.

### Technical Innovations and Methodological Insights

The consistent performance improvements achieved by MVE demonstrate the effectiveness of attention-based multi-view integration compared to simple averaging approaches. Unlike EchoPrime’s weighted integration resulting in high-dimensional representations (15 × 512 dimensions per study) ^3^, MVE enables dynamic weighting of echocardiographic views through self-attention mechanisms while compressing information into a single 512-dimensional study-level representation.

Importantly, while EchoPrime relies on explicit view classification and predefined weighting, MVE’s attention mechanism enables more fine-grained adaptive integration. This difference may be particularly effective for doppler echocardiography and non-routine views. Even within the same view category, color Doppler focusing on different blood flow regions, and zoom level variations cause substantial changes in focusing area. Such subtle variations cannot be captured by conventional view classification, but MVE’s attention mechanism can dynamically adjust importance based on the specific content and diagnostic value of each video. This flexibility enables appropriate integration between videos that provide different clinical information even from the same anatomical perspective, potentially contributing to improved model performance. This attention-based approach aligns with established clinical practice where experienced cardiologists routinely integrate information from multiple echocardiographic views to achieve accurate diagnoses ^1^.

### Fairness Challenges and Algorithmic Limitations

The limited effectiveness of adversarial learning in reducing demographic shortcuts through purely algorithmic interventions reveals fundamental challenges in achieving fairness. Systematic variation of adversarial weights from 0 to 1.0 showed that mild weighting still allowed access to demographic information, while excessive weighting substantially sacrificed downstream task performance. This trade-off suggests that relationships between demographic characteristics and echocardiographic features involve complex interactions between spurious correlations and legitimate biological differences that resist simple algorithmic disentanglement ^25^. Interestingly, with mild adversarial weighting (ω = 0.1), although demographic shortcut reduction was not evident, clinical outcome prediction performance slightly improved (Figure 2A). This suggests that adversarial learning may have learned more robust pathophysiological features that are less dependent on demographic shortcuts.

In addition, as shown in Figure 4, tasks with substantial subgroup disparities at baseline (ω = 0) (mild AS and LVEF<50%) showed greater vulnerability to performance degradation by adversarial weighting compared to tasks with minimal baseline disparities (impaired RV function). This differential response suggests that the degree of entanglement between demographic characteristics and pathophysiological features varies across cardiac evaluations. Thus, the persistence of demographic predictability in certain tasks may indicate appropriate preservation of clinically relevant biological information ^12,27^, rather than representing algorithmic failure.

### Clinical Implementation and Practical Considerations

For clinical translation, MVE requires users with basic understanding of machine learning workflows and VE concepts, combined with clinical expertise to assess echocardiographic video quality and interpret model outputs within appropriate clinical context. This level of expertise aligns with current expectations for AI-assisted diagnostic tools in cardiology practice ^28,29^, requiring clinical knowledge combined with basic AI literacy rather than specialized machine learning expertise.

Several data quality considerations must be addressed for successful implementation. Poor quality videos with excessive noise, inappropriate gain settings, or suboptimal imaging windows may result in unreliable VEs and should be flagged for manual review ^1,28^. These quality assurance measures are essential for maintaining diagnostic reliability in real-world clinical settings where image quality varies significantly from research datasets.

The efficiency gains achieved by our approach have important implications for sustainable AI development in healthcare. By reducing barriers from industrial-scale infrastructure to standard research computing resources, this framework has the potential to enable AI development in low-resource settings and developing healthcare systems ^10,11^. The findings suggest that developing sophisticated methods for extracting and refining representations from existing models may represent a more sustainable and accessible path toward advanced AI capabilities than continuously scaling model sizes.

### Limitations

Several important limitations should be considered when interpreting our findings. First, although our study utilized a substantial dataset (7,169 studies), the single-institution nature and temporal constraints (2017-2019) may limit generalizability across diverse healthcare systems, patient populations, and current clinical practice. However, external validation was limited by the lack of publicly available multi-view echocardiographic datasets, and this situation impedes the advancement of medical AI research. Future efforts toward public release of multi-view echocardiographic datasets by more institutions and implementation of multi-institutional validation studies will be essential for establishing broader generalizability. Second, dependence on EchoPrime foundation model embeddings creates potential dependencies on the quality and biases of the underlying foundation model. Any limitations or biases present in the foundation model may propagate through our framework, affecting downstream task performance and fairness assessments. Third, our MVE framework employs hyperparameters that require further optimization, including the fixed 512-dimensional embedding space and transformer architecture. The current dimensionality may be insufficient to capture all clinically relevant echocardiographic features, potentially limiting diagnostic performance for subtle or rare cardiac pathologies. Lastly, while our study focused on gradient reversal layer-based adversarial learning ^24^, other bias mitigation approaches such as disentangled representation learning or data reweighting methods ^30–32^ may be valuable for future investigation to determine their comparative effectiveness in echocardiographic analysis.

## Conclusion

MVE successfully democratizes access to advanced cardiac AI through efficient vector embedding approaches, enabling healthcare institutions with limited resources to achieve substantial diagnostic improvements without massive computational infrastructure. Notably, MVE VE showed robust performance with fewer echocardiographic views, demonstrating improved efficiency in extracting pathophysiological features—a critical advantage for real-world clinical implementation where comprehensive examinations are not always feasible.

While adversarial learning showed limited effectiveness in reducing demographic shortcuts, mild adversarial weighting (ω = 0.1) achieved the highest diagnostic performance improvements, suggesting potential benefits for learning more robust pathophysiological features. However, stronger adversarial weighting compromised diagnostic performance, revealing fundamental challenges in achieving algorithmic fairness.

This framework offers a practical pathway for broader AI adoption in cardiovascular medicine, though future research must address the complex interplay between diagnostic performance and demographic fairness in medical AI applications.

## Data Availability

The MIMIC-IV and MIMIC-IV-ECHO datasets analyzed in this study are publicly available through PhysioNet (https://physionet.org/) following completion of required training modules and approval of a data use agreement. The datasets used in this study cannot be directly redistributed due to patient privacy protections but can be obtained through the established PhysioNet access procedures.

## Acknowledgements

We thank the MIMIC-IV and MIMIC-IV-ECHO data contributors and the PhysioNet team for making these datasets publicly available.

## Code availability

Source code for MVE implementation, training procedures, and evaluation scripts are publicly available at “https://github.com/ttohya/mve-echo”.

