## supplement materials for "Multi-View Echocardiographic Embedding for Accessible AI Development"

### Supplementary Methods

#### Vector Embedding Preprocessing

**Vector Embedding Extraction for Each Video:** All echocardiographic videos were processed through the EchoPrime foundation model to generate 512-dimensional vector embeddings per video. Video preprocessing followed the EchoPrime procedures. Each study contained multiple videos (median: 41 videos, range: 1-118) including parasternal long-axis, short-axis, apical four-chamber, two-chamber, subcostal views, and color Doppler videos.

#### Demographic Data Processing for Adversarial Learning

**Sex:** Sex variable was processed with a three-category encoding scheme for female, male, and unknown/missing values. Missing values and non-standard entries were systematically mapped to the "unknown" category, enabling learning with all available data.

**Race:** Race variable underwent recategorization based on class imbalance: White and Black categories were maintained as separate classes, while other races including Hispanic and Asian were combined into an "Others" category.

#### Clinical Outcome Processing

**Binary task definition:** 21 Clinical outcomes were standardized into binary classification tasks encompassing echocardiographic measurements, functional assessments, and clinical diagnoses. Echocardiographic measurements utilized direct measurement data, with each task defined using guideline-based thresholds (e.g., LVEF < 50% for reduced ejection fraction, E/e' > 15 for elevated filling pressures).

**Clinical diagnosis labeling:** Clinical diagnoses were labeled using ICD codes and timestamps. For dilated cardiomyopathy and hypertrophic cardiomyopathy, patients with any recorded diagnosis were labeled as positive. For myocardial infarction and pulmonary embolism, a specific time window from 30 days before admission to 90 days post-discharge was set as a requirement for the diagnoses. Additionally, myocardial infarction cases required concurrent EF < 50% and pulmonary embolism cases required TRPG > 31 mmHg to ensure clinically significant cases and minimize false positive diagnoses. Negative cases were hospitalized patients who did not have these disease diagnoses based on ICD codes.

**ICD codes used for each clinical diagnosis:**

1. Dilated cardiomyopathy: 4254, I420
2. Hypertrophic cardiomyopathy: 4251*, I421, I422

3. Myocardial infarction candidates: 410*, 412, I21*, I22*, I252
4. Pulmonary embolism candidates: 415*, I26*

**Missing data handling:** Studies with missing outcome data for specific tasks were excluded from task-specific evaluations but retained for other available tasks. For shortcut learning assessment, unknown categories for sex and race were treated as missing data.

#### MVE and Adversarial Learning Model Architecture

**Input projection:** 512-dimensional vector embeddings from each echocardiographic view underwent linear projection, maintaining embedding dimensionality while enabling learnable feature transformation. To enable batch processing across studies with varying video counts, we implemented a fixed-length padding strategy with a maximum of 128 videos per study, covering all studies in MIMIC-IV-ECHO (maximum 118 videos per study).

**Shared transformer encoder architecture:** The encoder consisted of two transformer layers with eight attention heads per layer, 512-dimensional embeddings, and 2048-dimensional feedforward networks. The shared transformer simultaneously processed all available views within each study, enabling the model to learn inter-view dependencies through parallel processing.

**Masking and Aggregation Strategy:** Two masking approaches were implemented: padding masks to ignore zero-padded positions during attention computation, and 50% random masking of view embeddings during pre-training using learnable mask tokens. A global query mechanism aggregated valid views through multi-head attention to create study-level representations, with reconstruction performed using a linear layer recovering original 512-dimensional embeddings.

**Adversarial discriminator architecture:** The adversarial component employed gradient reversal layers to minimize demographic predictability. The discriminator included separate classification heads for sex prediction (3 classes) and race prediction (4 classes). The discriminator consisted of feedforward networks (512→3 dimensions and 512→4 dimensions) for demographic prediction.

#### Training Implementation

**Loss function implementation:** The training objective combined reconstruction loss and adversarial loss components (L_total = L_reconstruction + ω × L_adversarial). Reconstruction loss used mean squared error calculated only for masked view positions. Adversarial loss combined separate cross-entropy losses for sex and race prediction.

**Optimization strategy:** Training used the AdamW optimizer with learning rate $1\times10^{-4}$ and weight decay 0.01. Pre-training consisted of 50 epochs with fixed learning rate scheduling and batch size of 8 studies per iteration.

**Computational Efficiency:** All experiments were conducted on a single NVIDIA RTX A6000 GPU. MVE training completed in under 10 minutes, demonstrating significant efficiency compared to foundation model training requiring weeks to months. The primary bottleneck was vector embedding extraction with the foundation model (EchoPrime), converting 1.6TB of DICOM data to 2.7GB of embeddings over 2-3 days, but this one-time preprocessing enables rapid subsequent model development.

#### Evaluation Implementation

**Subject-based cross-validation implementation:** Cross-validation used GroupKFold with four folds to strictly maintain subject-level independence. The splitting algorithm first identified unique subjects, then assigned subjects and their associated studies to different folds to ensure complete population separation.

**Multi-metric performance evaluation:** All downstream tasks employed logistic regression with L2 regularization. The L2 regularization parameter was adjusted proportionally to embedding dimensionality (base $\lambda = 0.01 \times 512/\text{input\_dim}$) to ensure consistent regularization strength across different representation sizes. Threshold setting used values that maximized F1 scores on the training set. Performance evaluation adopted area under the ROC curve (AUC) as the primary metric, complemented by accuracy and F1 scores.

**Demographic predictability evaluation:** Bias assessment used separate classification tasks for sex (2 classes: F/M, excluding unknown) and race (3 classes: White/Black/Other, excluding unknown). Demographic predictability was evaluated using the same cross-validation framework as downstream tasks with identical logistic regression configuration.

#### Statistical Testing Details

**Performance analysis and shortcut evaluation:** We implemented statistical analysis using mixed effects models for MVE VE performance and fairness evaluation. This analysis compared performance between Foundation VE (baseline model) and MVE VEs with different adversarial weightings, specifically evaluating predictive performance improvement in clinical tasks and fairness improvement for demographic attributes (sex and race). The mixed effects models treated each repeat-fold combination as a separate observation, with random effects accounting for variability between these combinations.

For performance analysis, we used the following mixed effects model:

$$\boldsymbol{AUC\_ijkl = \beta_0 + \beta_1(model\_type\_i) + u\_jk + v\_l + \varepsilon\_ijkl}$$

where AUC_ijkl represents the performance for model type i, repeat j, fold k, and task l; model_type was treated as a fixed effect; u_jk represents random intercepts for independent repeat-fold combinations; v_l represents random intercepts for clinical tasks; and ε_ijkl represents residual error. This allows appropriate adjustment for variability between independent repeat-fold combinations and different clinical tasks.

For fairness analysis, we used the following mixed effects model:

$$\boldsymbol{AUC\_ijk = \beta_0 + \beta_1(model\_type\_i) + u\_jk + \varepsilon\_ijk}$$

where AUC_ijk represents demographic prediction performance for model type i, repeat j, and fold k; model_type_i was treated as a fixed effect; u_jk represents random intercepts for independent repeat-fold combinations; and ε_ijk represents residual error. This approach evaluates demographic shortcut learning, where lower AUC values in predicting demographic attributes from learned representations indicate reduced reliance on demographic shortcuts and improved fairness.

To control for multiple comparisons across models with different adversarial weightings, we applied the Holm-Bonferroni method. This enables detection of true improvement effects while appropriately controlling false positive rates, calculating 95% confidence intervals and corrected p-values for each comparison.

**Bias Evaluation among VE types:** We implemented statistical methods to evaluate subgroup performance differences using mixed effects models in MVE analysis. This approach quantitatively assessed performance differences across subgroups defined by patient characteristics including sex (male/female), race (white/black/other), and video count (<20, 20-39, 40-59, ≥60) for different models (Foundation VE, MVE VE and its adversarial weight variations).

We used the following mixed effects model:

$$\begin{aligned}\boldsymbol{AUC\_ijklm} &= \boldsymbol{\beta_0 + \beta_1(model\_type\_i) + \beta_2(subgroup\_j)} \\ &\quad \boldsymbol{+ \beta_3(model\_type\_i \times subgroup\_j) + u\_kl + v\_m + \varepsilon\_ijklm}\end{aligned}$$

where AUC_ijklm represents performance for model type i, subgroup j, repeat k, fold l, and task m; model_type_i and subgroup_j were treated as fixed effects; model_type_i × subgroup_j represents the interaction term; u_kl represents random intercepts for independent repeat-fold combinations; v_m represents random intercepts for clinical tasks; and ε_ijklm represents residual error. This design enables simultaneous testing of overall performance differences between models, systematic differences between subgroups, differences in improvement effects between subgroups (interaction effects). No multiple testing correction was applied for this analysis.

**Supplementary Figure 1: Comprehensive data flow diagram of the multi-view encoder framework**

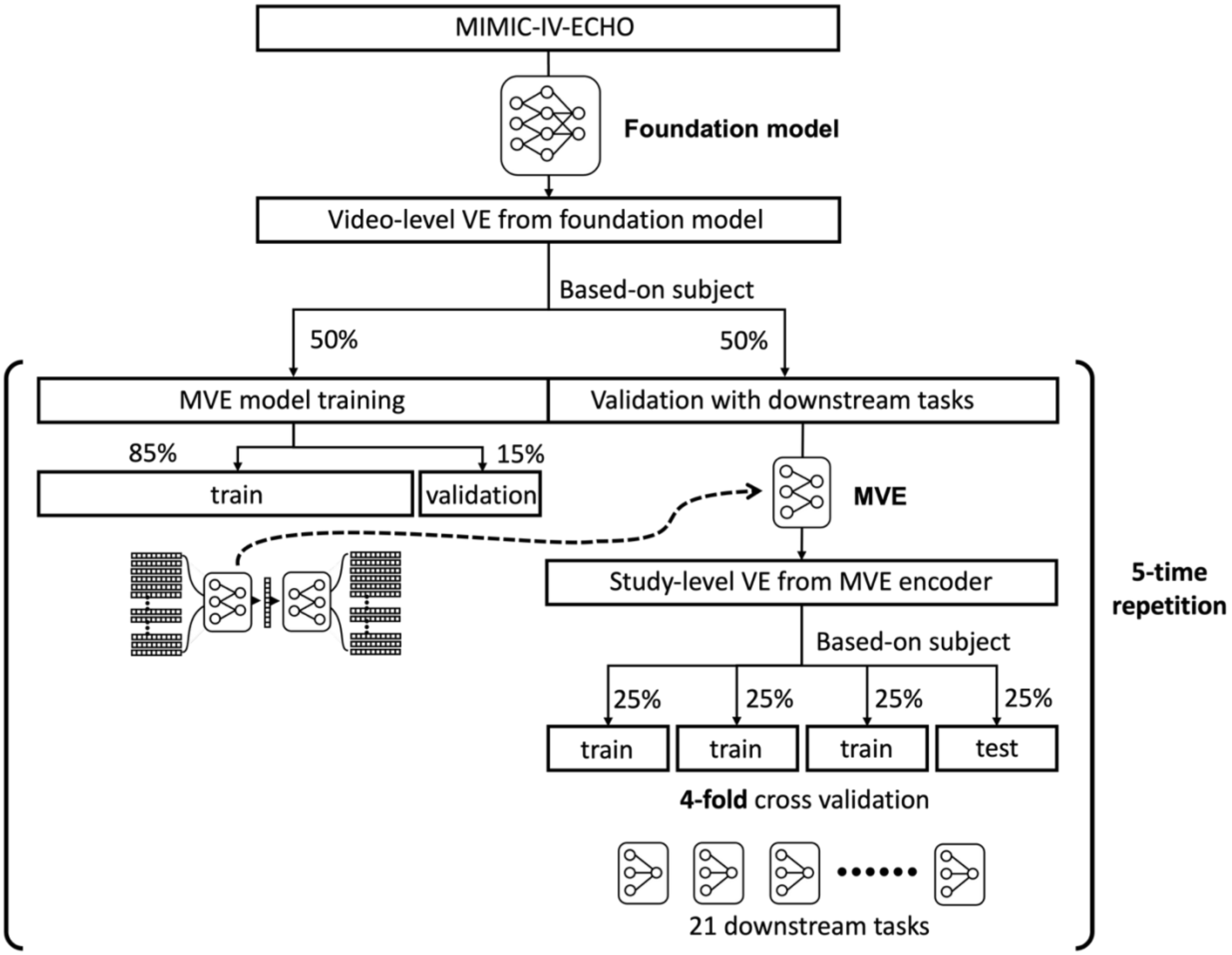


Complete experimental pipeline showing a multi-view encoder (MVE) implementation. All DICOM video files were converted to video-level vector embeddings (VE) using the EchoPrime foundation model. VE data were split into training and validation sets (50:50) using subject-based splitting to prevent information leakage. The MVE was trained using the training dataset with adversarial learning components. Validation data were converted to study-level VE using the trained MVE. These study-level embeddings were used to evaluate 21 downstream cardiac diagnostic tasks and shortcut learning for sex and race prediction through 4-fold cross-validation. This entire process was repeated five times using different random seeds for robust evaluation.

**Supplementary Figure 2: Comprehensive performance metrics across all tasks**

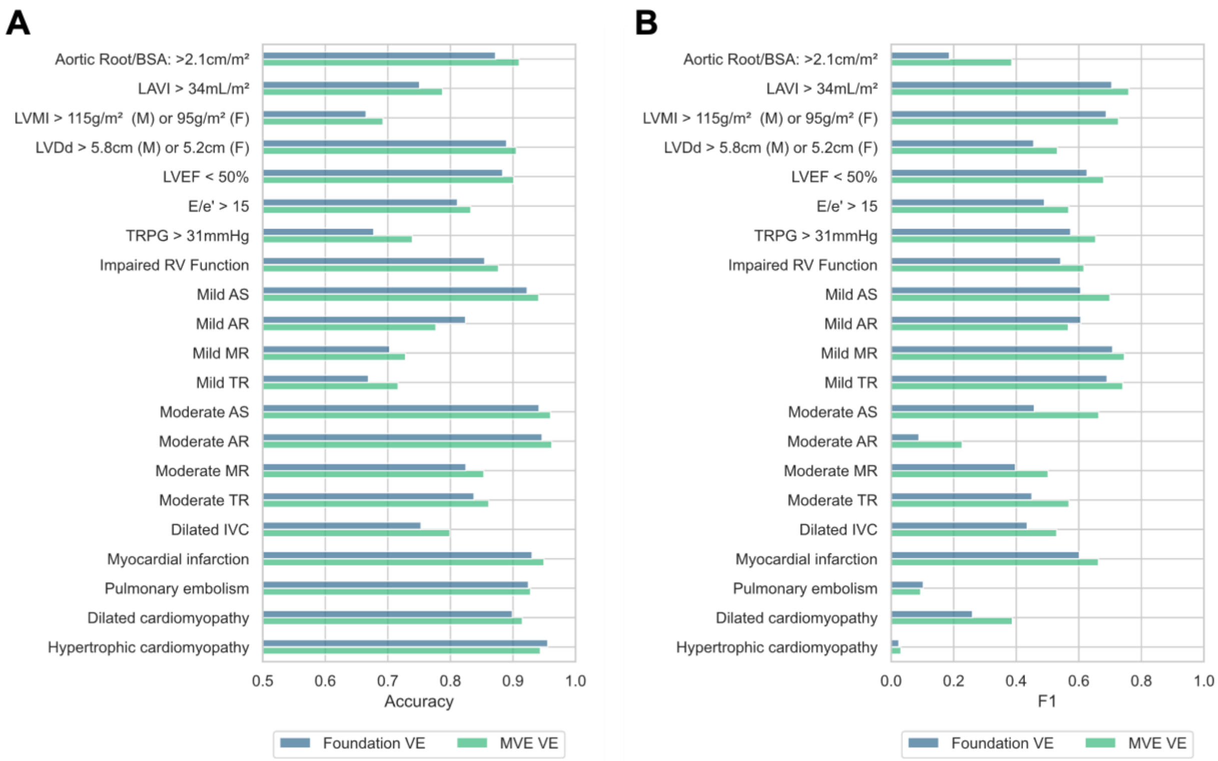


Detailed comparison of accuracy (A) and F1 score (B) performance between vector embeddings from foundation model (Foundation VE) and multi-view encoder without adversarial training (MVE VE, ω = 0.1) for all 21 cardiovascular evaluation tasks.

**sTable 1: MIMIC-IV-ECHO dataset characteristics**

| Variables | Missing | Overall 7169 | Female 3478 | Male 3274 | White 4777 | Black 1232 | Others 662 |
|---|---|---|---|---|---|---|---|
| Mild AS or greater, n (%) | 171 | 726 (10.4) | 361 (10.5) | 331 (10.3) | 571 (12.2) | 67 (5.5) | 49 (7.5) |
| LVEF < 50%, n (%) | 1804 | 830 (15.5) | 321 (11.9) | 493 (20.9) | 554 (15.5) | 184 (19.7) | 70 (14.1) |
| Impaired RV Function, n (%) | 450 | 1064 (15.8) | 424 (12.9) | 629 (20.4) | 721 (16.0) | 223 (19.3) | 103 (16.3) |
| E/e' > 15, n (%) | 4064 | 549 (17.7) | 326 (22.0) | 216 (15.3) | 382 (18.5) | 102 (19.5) | 55 (19.5) |

AS: Aortic Stenosis, LVEF: Left Ventricular Ejection Fraction, RV: Right Ventricle, E/e': Ratio of early mitral inflow velocity to early diastolic mitral annular tissue velocity.
